# Effects of long-term unmet needs and unmet rehabilitation needs on the quality of life in stroke survivors

**DOI:** 10.1101/2024.03.08.24304010

**Authors:** Yookyung Lee, Won-Seok Kim, Won Kee Chang, Yun Sun Jung, Sungju Jee, Sung-Hwa Ko, Min Kyun Sohn, Yong-Il Shin, Hee-Joon Bae, Beom Joon Kim, Jun Yup Kim, Dong-Ick Shin, Kyu Sun Yum, Hee-Yun Chae, Dae-Hyun Kim, Jae-Kwan Cha, Man-Seok Park, Joon-Tae Kim, Kang-Ho Choi, Jihoon Kang, Nam-Jong Paik

**Author notes:** **Corresponding authors** Nam-Jong Paik, MD, PhD, Department of Rehabilitation Medicine, Seoul National University College of Medicine, Seoul National University Bundang Hospital, 82 Gumi-ro 173 Beon-gil, Bundang-gu, Seongnam 13620, Korea, Jihoon Kang, MD, PhD, Department of Neurology, Cerebrovascular Center, Seoul National University Bundang Hospital, Seoul National University, 82 Gumi-ro 173 Beon-gil, Bundang-gu, Seongnam 13620, Korea. These authors contributed equally to this work (Yookyung Lee, Won-Seok Kim).

## Abstract

**Background:** Unmet long-term needs and rehabilitation needs are prevalent among stroke survivors and affect their quality of life. We aimed to identify the long-term unmet needs and unmet rehabilitation needs among stroke survivors in South Korea and evaluate their intercorrelations with health-related quality of life.

**Methods:** Stroke survivors who were admitted to four Regional Cardiocerebrovascular Disease Centers between January 1, 2015 and December 31, 2019 were telephonically surveyed using a computer-assisted telephone interview method. With the aim of surveying approximately 1,000 patients, 9,204 people were recruited through random sampling. Unmet needs were evaluated on the basis of Longer-term Unmet Needs after Stroke questionnaire items. Quality of life was evaluated using the EuroQoL 5-dimension, 3-level (EQ-5D-3L) questionnaire and the EQ-5D index.

**Results:** Among the participants, 93.6% experienced at least one unmet need and 311 (32.6%) reported unmet rehabilitation needs. The number of unmet needs, age, modified Rankin Scale (mRS) score, and previous stroke showed significant negative correlations with the EQ-5D index (p-value < 0.05). The age-adjusted odds ratio (OR) for reporting unmet rehabilitation needs significantly increased with problems in mobility (OR, 4.96; 95% confidence interval [CI], 3.64-6.76), self-care (OR, 4.46; 95% CI, 3.32-5.98), usual activities (OR, 5.78; 95% CI, 4.21-7.93), pain/discomfort (OR, 3.76; 95% CI, 2.76-5.06), anxiety/depression (OR, 3.67; 95% CI, 2.74-4.91), higher mRS score (OR, 3.13; 95% CI, 2.29-4.28), prior hyperlipidemia (OR, 1.35; 95% CI, 1.00-1.81), and number of unmet needs (OR, 1.30; 95% CI, 1.25-1.36).

**Conclusions:** Unmet needs were prevalent among stroke survivors and were associated with a lower quality of life and increased odds of reporting unmet rehabilitation needs. Further research is needed to investigate strategies for addressing these subjective unmet needs with the aim of improving the long-term quality of life of stroke survivors.

## Introduction

Stroke is the second-leading cause of death and the third-leading cause of disability worldwide (1). It represents a substantial global burden, and its incidence is expected to increase with age (2,3). Many stroke survivors have disabilities, and unmet needs are highly prevalent among them (4,5). Depending on the study population and measurement tools, up to 97.5% of stroke survivors have reported long-term unmet needs after stroke (4). The presence of unmet long-term needs has been closely associated with poor functional outcomes (6,7).

Stroke survivors frequently experience unmet rehabilitation needs. Approximately one in three patients reported unmet rehabilitation needs post-stroke (7). These needs can influence the patients’ health-related quality of life (7,8). A previous study showed that in community-dwelling stroke survivors, an increased number of unmet rehabilitation needs was significantly associated with a lower health-related quality of life (8).

Although numerous studies have explored long-term unmet needs and rehabilitation needs following stroke, the majority of these investigations have been conducted in Western countries. Studies of the West Pacific population are sparse and characterized by small sample sizes (4,5). The factors associated with unmet needs and their impact on outcomes and health-related quality of life remain largely unexplored within these populations. Consequently, we aimed to identify the long-term unmet needs and unmet rehabilitation needs among stroke survivors in South Korea and evaluate their intercorrelations with health-related quality of life.

## Methods

This cross-sectional study followed the strengthening the reporting of observational studies in epidemiology guideline (9).

### Data Source

In South Korea, Regional Cardiocerebrovascular Disease Centers (RCCVCs) were designated by the Ministry of Health and Welfare (https://www.mohw.go.kr/eng/) in 2008 for better prevention and treatment of stroke and acute myocardial infarction (10). Currently, 14 hospitals are designated as RCCVCs nationwide. The RCCVC registry for acute ischemic stroke was launched in 2014, and efforts to improve data entry and prevent errors have been made through regular registry committee meetings and training workshops.

### Study Participants

For this study, using the RCCVC registry databases of four centers (Seoul National University Bundang Hospital, Dong-A University Hospital, Jeonnam National University Hospital, and Chungbuk National University Hospital), we identified stroke survivors who were admitted to these centers between January 1, 2015, and December 31, 2019. Patients who provided consent for the use of their contact information, were aged 18 years or older, and had a modified Rankin Scale (mRS) score of 1-5 at 3 months post-stroke were included. Patients who were not community-dwelling or died within one year of follow-up were excluded. The number of eligible patients extracted from the databases of the four RCCVCs for the telephone survey was 9,204. The telephone survey was conducted by trained interviewers in Gallup Korea using a computer-assisted telephone interview method from December 2, 2020, to December 14, 2020. To survey 1,000 patients, 9,204 were recruited through random sampling. Among these 9,204 patients, 6,022 could not be reached and dropped out of the study. Of the 3,182 people contacted, 1,419 refused to participate, were busy, absent, or gave inappropriate responses and dropped out. A total of 761 patients were excluded because of death or admission to facilities such as hospitals and nursing homes. In total, 1,002 survey respondents were included in the final analysis (Figure 1).

**Figure 1.**
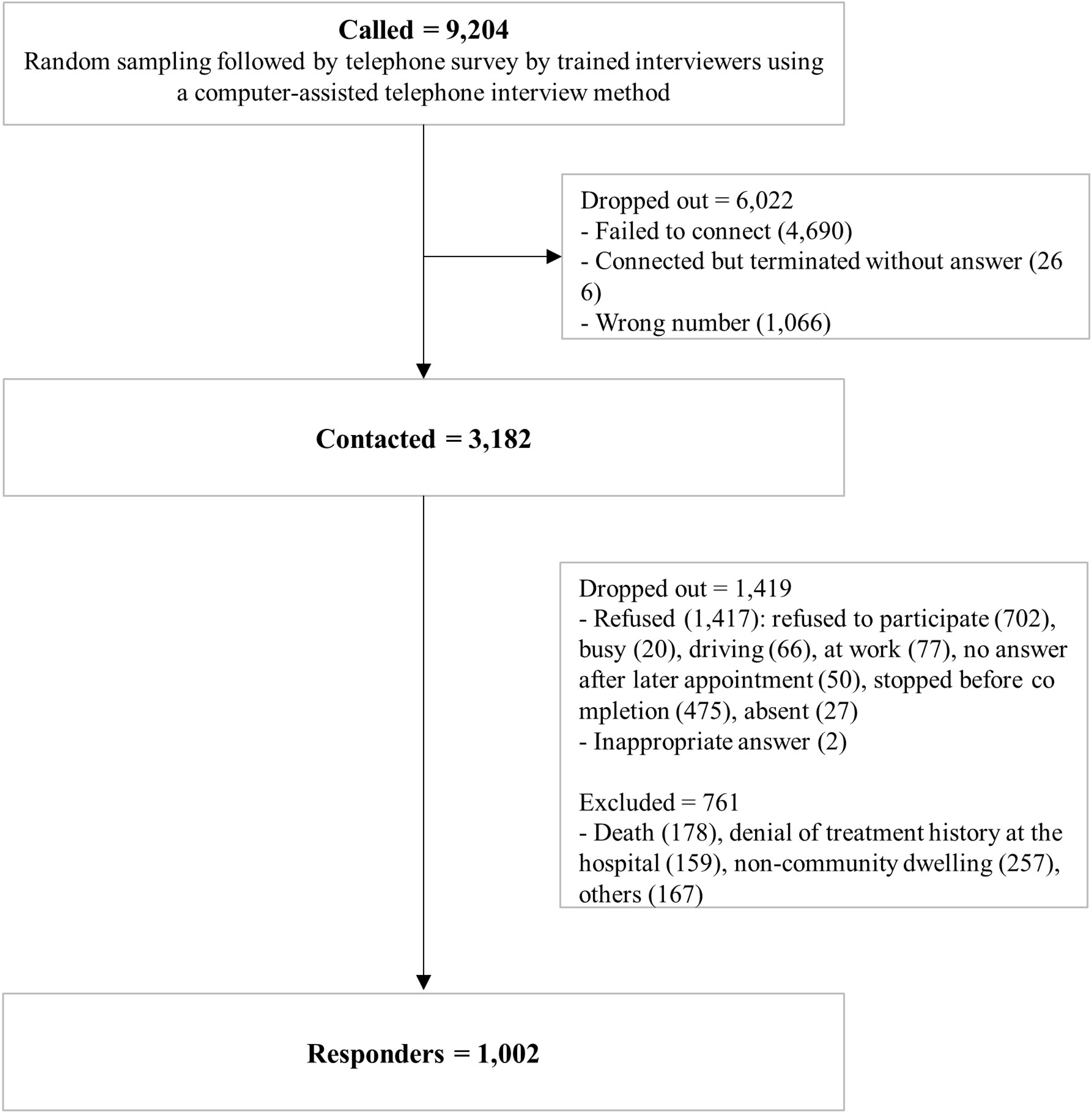
Study flow

### Telephone Survey Questionnaire

Screening was performed using the following questions: 1) Have you received stroke treatment at OOO Hospital? 2) Are you currently staying in a hospital, nursing hospital, nursing home, or another facility? If the answer was “no” for question 1 and “yes” or “deceased” for question 2, the patient was excluded, and the survey was terminated.

Unmet needs were evaluated using the Longer-term Unmet Needs after Stroke (LUNS) questionnaire items, which were developed to measure survivors’ long-term physical, social, and emotional well-being after stroke (11). The LUNS was translated and adapted to Korean and validated in 66 adults with stroke in our previous study (12). Considering the sentiments of Korean seniors, who tend to avoid discussing physical relationships, we excluded the “Advice on physical relationships” item from the 22 items of the LUNS. Quality of life (QoL) was evaluated using the EuroQoL 5-dimension, 3-level (EQ-5D-3L) questionnaire, and the EQ-5D index ranging from 0 (death) to 1 (best) was calculated using the Korean valuation set (13). Current functional status was assessed using a simplified modified Rankin Scale (mRS) questionnaire (14), and socioeconomic data, including the participants’ education and residential area and whether they were living alone or not, were also collected. The perceived adequacy of rehabilitation after stroke was examined using the following question: Do you think that rehabilitation after stroke was adequately delivered? The response options were (i) completely inadequate; (ii) inadequate; (iii) adequate; (iv) very adequate; and (v) no need for rehabilitation. Responses 1 and 2 indicated perceived unmet rehabilitation needs, while responses 3-5 were considered to indicate met rehabilitation needs.

### Stroke-Related Variables

Stroke-related variables, such as stroke type, initial National Institutes of Health Stroke Scale (NIHSS) score, discharge status, and demographic information, were gathered from the stroke registries of the RCCVCs. Medical histories associated with stroke risk, such as previous stroke, transient ischemic attack, coronary heart disease, hypertension, diabetes mellitus, hyperlipidemia, and atrial fibrillation, were also collected.

### Statistical Analysis

All categorical values were presented as n (%), and the total number of unmet needs and EQ-5D scores were presented as mean ± standard deviation. Multiple linear regression analysis with backward elimination was performed to analyze the factors associated with health-related QoL as measured using the EQ-5D. The t-test and chi-square test were used to analyze perceived unmet rehabilitation needs in relation to participant characteristics. Multiple logistic regression analysis was used to analyze factors associated with perceived unmet rehabilitation needs.

All statistical analyses were performed using SPSS for Windows software package (ver. 22.0; SPSS Inc., Chicago, IL, USA). Statistical significance was set at P < 0.05.

### Ethics Statement

This study was conducted in accordance with the Declaration of Helsinki. The study protocol was approved by the Institutional Review Board of Seoul National University Bundang Hospital (IRB No. B-2009-639-302) and Chungbuk National University Hospital (IRB No. 2020-12-014).

## Results

The baseline characteristics of the study participants (n = 1,002) are shown in Table 1. The mean participant age was 69.26 ± 12.43 years, and 635 (63.4%) participants were men. The prevalence of participants with at least one unmet need was 93.6%, and the mean number of unmet needs was 4.67 ± 3.65. A total of 311 (32.6%) individuals stated that their rehabilitation needs were unmet.

**Table 1.**
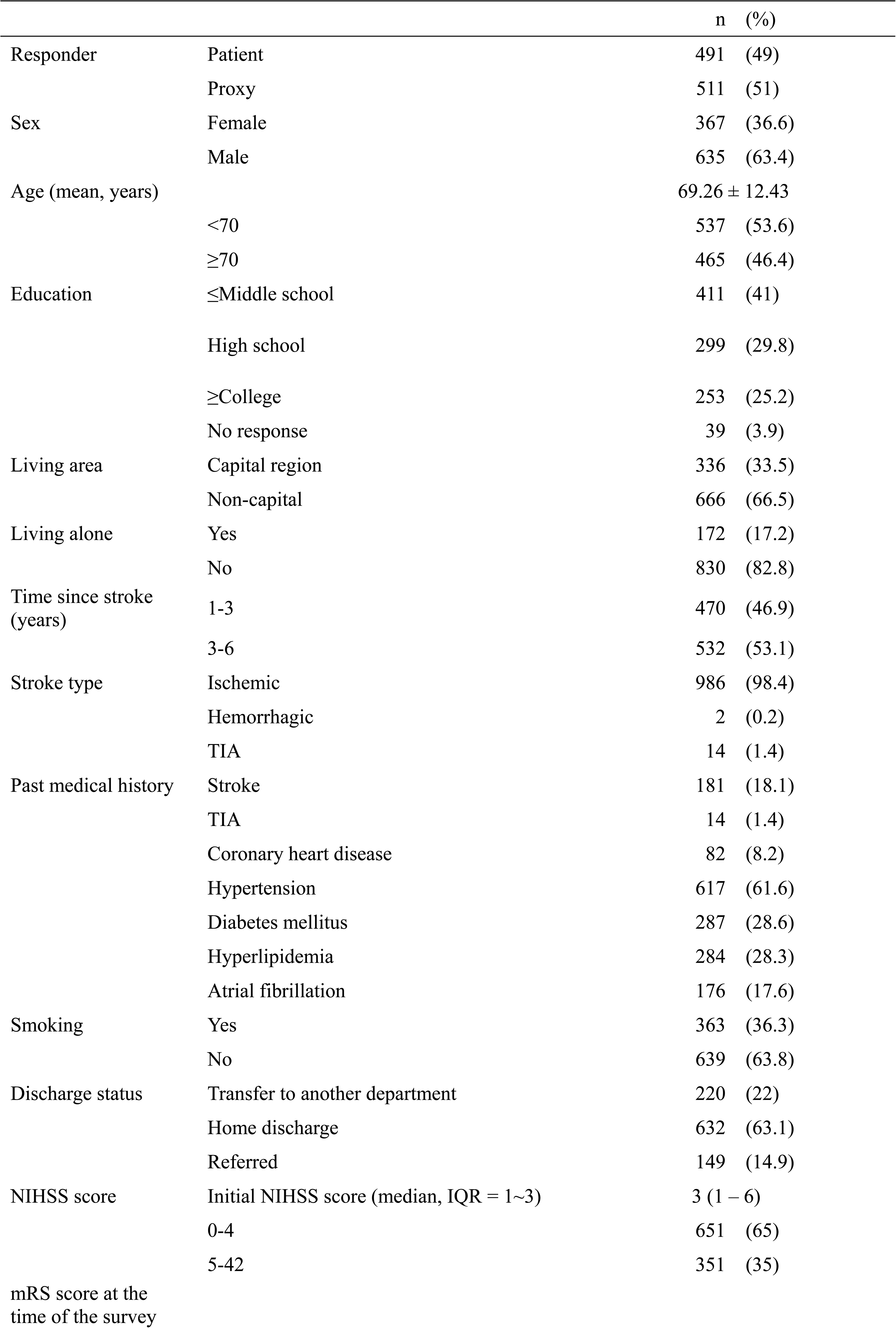
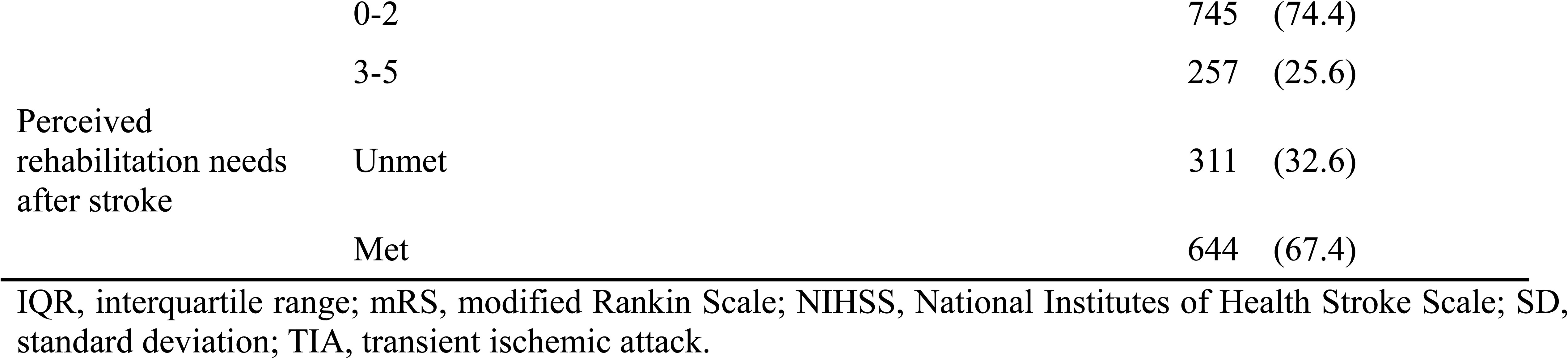
Baseline characteristics of the participants (N = 1,002)

|  |  | n | (%) |
| --- | --- | --- | --- |
| Responder | Patient | 491 | (49) |
|  | Proxy | 511 | (51) |
| Sex | Female | 367 | (36.6) |
|  | Male | 635 | (63.4) |
| Age (mean, years) |  | 69.26 ± 12.43 |  |
|  | <70 | 537 | (53.6) |
|  | ≥70 | 465 | (46.4) |
| Education | ≤Middle school | 411 | (41) |
|  | High school | 299 | (29.8) |
|  | ≥College | 253 | (25.2) |
|  | No response | 39 | (3.9) |
| Living area | Capital region | 336 | (33.5) |
|  | Non-capital | 666 | (66.5) |
| Living alone | Yes | 172 | (17.2) |
|  | No | 830 | (82.8) |
| Time since stroke (years) | 1-3 | 470 | (46.9) |
|  | 3-6 | 532 | (53.1) |
| Stroke type | Ischemic | 986 | (98.4) |
|  | Hemorrhagic | 2 | (0.2) |
|  | TIA | 14 | (1.4) |
| Past medical history | Stroke | 181 | (18.1) |
|  | TIA | 14 | (1.4) |
|  | Coronary heart disease | 82 | (8.2) |
|  | Hypertension | 617 | (61.6) |
|  | Diabetes mellitus | 287 | (28.6) |
|  | Hyperlipidemia | 284 | (28.3) |
|  | Atrial fibrillation | 176 | (17.6) |
| Smoking | Yes | 363 | (36.3) |
|  | No | 639 | (63.8) |
| Discharge status | Transfer to another department | 220 | (22) |
|  | Home discharge | 632 | (63.1) |
|  | Referred | 149 | (14.9) |
| NIHSS score | Initial NIHSS score (median, IQR = 1~3) | 3 | (1 – 6) |
|  | 0-4 | 651 | (65) |
|  | 5-42 | 351 | (35) |
| mRS score at the time of the survey |  |  |  |

|  |  |  |
| --- | --- | --- |
|  | 0-2 | 745 (74.4) |
|  | 3-5 | 257 (25.6) |
| Perceived<br>rehabilitation needs<br>after stroke | Unmet | 311 (32.6) |
|  | Met | 644 (67.4) |

IQR, interquartile range; mRS, modified Rankin Scale; NIHSS, National Institutes of Health Stroke Scale; SD, standard deviation; TIA, transient ischemic attack.

The most frequently cited unmet needs after stroke were “Help with applying for benefits” (48.4%), “Advice on employment” (46.6%), and “Fear of falling” (37.7%) (Supplementary table 1). In the multiple linear regression analysis with backward elimination, the number of unmet needs, perceived unmet rehabilitation needs, age, mRS score, and previous stroke were significant factors that showed negative correlations with the EQ-5D index (Table 2). When adjusted for age, sex, and mRS score at the time of the survey, the total number of unmet needs differed significantly according to living area, education level, living alone, problems with mobility, self-care, usual activities, pain/discomfort, and anxiety/depression (Supplementary table 2).

**Table 2.** Multiple linear regression model for health-related quality of life measured by the EQ-5D index.

| Variables | $\beta$ | SE | p-value |
| --- | --- | --- | --- |
| Number of unmet needs | -0.184 | 0.002 | <0.001 |
| Perceived unmet rehabilitation needs | -0.075 | 0.013 | 0.001 |
| Age | -0.049 | 0 | 0.018 |
| Modified Rankin Scale score | -0.650 | 0.004 | <0.001 |
| Previous stroke | -0.061 | 0.014 | 0.003 |

SE, standard error.
F = 327.888 (p < 0.001); adjusted $R^2$ for model = 0.650; Durbin-Watson = 2.132

Those reporting unmet rehabilitation needs were significantly older; had lower education levels; experienced problems in mobility, self-care, usual activities, pain/discomfort, anxiety/depression; and showed a higher mRS score at the time of survey, higher initial NIHSS score, prior stroke, higher number of unmet needs, and lower EQ-5D index values (Table 3).

**Table 3.** Perceived unmet rehabilitation needs after stroke.

|  |  | Unmet<br>n = 311 (%) | Met<br>n = 644 (%) | Total<br>n = 955 (%) | p-value |
| --- | --- | --- | --- | --- | --- |
| Age (years)† | (mean ± SD) | 70.6±11.5 | 68.4±12.8 | 69.1±12.4 | 0.008 |
|  | <70 | 119 (38.3) | 314 (48.8) | 433 (45.3) | 0.002 |
|  | ≥70 | 192 (61.7) | 330 (51.2) | 522 (54.7) |  |
| Sex | Female | 119 (38.3) | 229 (35.6) | 348 (36.4) | 0.430 |
|  | Male | 192 (61.7) | 415 (64.4) | 607 (63.6) |  |
| Area of residence | Capital region | 96 (30.9) | 227 (35.2) | 323 (33.8) | 0.189 |
|  | Non-capital | 215 (69.1) | 417 (64.8) | 632 (66.2) |  |
| Education* | ≤Middle school | 141 (46.5) | 241 (39.0) | 382 (41.5) | 0.011 |
|  | High school | 99 (32.7) | 193 (31.2) | 292 (31.7) |  |
|  | ≥College | 63 (20.8) | 184 (29.8) | 247 (26.8) |  |
| Living alone | Yes | 53 (17.0) | 108 (16.8) | 161 (16.9) | 0.927 |
|  | No | 258 (83.0) | 536 (83.2) | 794 (83.1) |  |
| Time since stroke (years) | 1-3 | 159 (51.1) | 293 (45.5) | 452 (47.3) | 0.112 |
|  | 3-6 | 152 (48.9) | 351 (54.5) | 503 (52.7) |  |
| Problems in mobility† | No | 79 (25.4) | 405 (62.9) | 484 (50.7) | <0.001 |
|  | Yes | 232 (74.6) | 239 (37.1) | 471 (49.3) |  |
| Problems in self-care† | No | 119 (38.3) | 475 (73.8) | 594 (62.2) | <0.001 |
|  | Yes | 192 (61.7) | 169 (26.2) | 361 (37.8) |  |
| Problems in usual activities† | No | 68 (21.9) | 400 (62.1) | 468 (49.0) | <0.001 |
|  | Yes | 243 (78.1) | 244 (37.9) | 487 (51.0) |  |
| Problems in pain/discomfort† | No | 84 (27.5) | 371 (58.6) | 455 (48.5) | <0.001 |
|  | Yes | 221 (72.5) | 262 (41.4) | 483 (51.5) |  |
| Problems in anxiety/depression† | No | 99 (33.1) | 408 (64.9) | 507 (54.6) | <0.001 |
|  | Yes | 200 (66.9) | 221 (35.1) | 421 (45.4) |  |
| mRS score at the time of the survey† |  |  |  |  | <0.001 |
|  | 0-2 | 184 (59.2) | 531 (82.5) | 715 (74.9) | <0.001 |
|  | 3-5 | 127 (40.8) | 113 (17.5) | 240 (25.1) |  |
| Initial NIHSS score |  | 5.2 ± 5.0 | 4.4 ± 4.8 |  | 0.015 |
|  | 0-4 | 186 (59.8) | 436 (67.7) |  | 0.017 |
|  | 5-42 | 125 (40.2) | 208 (32.3) |  |  |
| Prior stroke* | No | 244 (78.5) | 540 (83.9) | 784 (82.1) | 0.047 |
|  | Yes | 67 (21.5) | 104 (16.1) | 171 (17.9) |  |
| Prior hyperlipidemia | No | 209 (67.2) | 472 (73.3) | 681 (71.3) | 0.056 |
|  | Yes | 102 (32.8) | 172 (26.7) | 274 (28.7) |  |
| Number of unmet needs† | (mean ± SD) | 6.9 ± 3.8 | 3.6 ± 3.0 | 4.6 ± 3.7 | <0.001 |

mRS, modified Rankin Scale; NIHSS, National Institutes of Health Stroke Scale; SD, standard deviation.
\*p < 0.05, †p < 0.01

The age-adjusted odds ratio (OR) for reporting unmet rehabilitation needs significantly increased with problems in mobility (OR, 4.96; 95% confidence interval [CI], 3.64-6.76), self-care (OR, 4.46; 95% CI, 3.32-5.98), usual activities (OR, 5.78; 95% CI, 4.21-7.93), pain/discomfort (OR, 3.76; 95% CI, 2.76-5.06), anxiety/depression (OR, 3.67; 95% CI, 2.74-4.91), higher mRS score (OR, 3.13; 95% CI, 2.29-4.28), prior hyperlipidemia (OR, 1.35; 95% CI, 1.00-1.81), and number of unmet needs (OR, 1.30; 95% CI, 1.25-1.36) (Table 4). The OR significantly decreased with an educational level of college or higher (OR, 0.66; 95% CI, 0.45-0.96). In the final model of multiple logistic regression analysis with backward elimination, problems in mobility (OR, 1.78; 95% CI, 1.16-2.74), usual activities (OR 1.95; 95% CI 1.24-3.06), and higher number of unmet needs (OR 1.22; 95% CI 1.16-1.28) showed significant associations with perceived unmet rehabilitation needs (Supplementary table 3).

**Table 4.** Age-adjusted logistic regression for perceived unmet rehabilitation needs.

| Variable | OR | 95% CI |  |
| --- | --- | --- | --- |
|  |  | Lower | Upper |
| Education level college or higher* | 0.66 | 0.45 | 0.96 |
| Problems in mobility† | 4.96 | 3.64 | 6.76 |
| Problems in self-care† | 4.46 | 3.32 | 5.98 |
| Problems in usual activities† | 5.78 | 4.21 | 7.93 |
| Problems in pain/discomfort† | 3.76 | 2.79 | 5.06 |
| Problems in anxiety/depression† | 3.67 | 2.74 | 4.91 |
| Modified Rankin Scale score† | 3.13 | 2.29 | 4.28 |
| Prior hyperlipidemia* | 1.35 | 1.00 | 1.81 |
| Number of unmet needs† | 1.30 | 1.25 | 1.36 |

CI, confidence interval; OR, odds ratio.
† p < 0.01
\* p < 0.05

## Discussion

Unmet and perceived unmet rehabilitation needs are highly prevalent among community-dwelling stroke survivors. In the present study, the number of unmet and perceived unmet rehabilitation needs was found to have a detrimental impact on health-related QoL, as assessed using the EQ-5D index. The likelihood of perceived unmet rehabilitation needs increased with an increase in the number of unmet needs and problems in the EQ-5D dimensions. Educational level, mRS score, and prior hyperlipidemia were also associated with perceived unmet rehabilitation needs.

### Long-term Unmet Needs after Stroke

Most stroke survivors in the present study (93.6%) had long-term unmet needs. A recent systematic review reported that the prevalence of unmet needs among stroke survivors ranged from 15% to 97.5% (4). The prevalence of unmet needs in our study was at the higher end of this range. In our study, we used the LUNS questionnaire to evaluate unmet needs. Two previous studies using this questionnaire reported prevalence rates of 67.7% and 85.1%, respectively (11,15). The time post-stroke was 5-8 years in the one study and 3–6 months in the other. Our study evaluated stroke survivors 1-6 years post-stroke. The subtle disparities in prevalence observed among these studies may be attributed to variations in post-stroke duration, the compositions of the study populations, and the influence of social and cultural factors (5).

In our study, the most frequently cited unmet needs were “Help with applying for benefits,” “Advice on employment,” and “Fear of falling.” These unmet needs contrast with those identified in a prior Dutch study of stroke survivors 5-8 years after stroke. In that study, which also used the LUNS questionnaire, “Information on stroke” was the most frequently mentioned unmet need followed by “Fear of falling” and “Help with concentration or memory” (15). In Korea, benefits for stroke survivors include the National Disability Registration System (NDRS) and long-term care (LTC) for the elderly (16,17). Stroke survivors are eligible for disability registration 6 months after the stroke (17). In 2008, Korea introduced LTC insurance to cover older adults in need of LTC. People aged 65 years or older or under 65 years with geriatric diseases (such as dementia cerebrovascular disease, Parkinson disease, and other neurological diseases) are eligible for LTC services (16). The unmet need in “Help with applying for benefits” may be due to lack of knowledge about the available benefits or difficulties in the process of applying for benefits. Thus, information on applying for benefits for persons with disabilities should be communicated in a manner that considers the specific challenges of stroke survivors, including limited transportation, issues related to digital literacy, communication problems, and cognitive impairments.

“Advice on employment” was an unmet need reported by 46.6% of stroke survivors. A previous study by Chang et al. reported that the rate of return to work was 60% in functionally independent Korean stroke survivors 6 months post-stroke (18). Older female stroke survivors with comorbidities and lower educational levels were less likely to return to work (18). Interventions to facilitate return to work include multidisciplinary assessment, identification of barriers to work, evaluation of the working environment, communication with employers, rehabilitation of working skills, education, and counseling (19). However, standardized rehabilitation programs are unavailable, and evidence of the effectiveness of interventions remains weak (19). In this regard, additional research and implementation of vocational rehabilitation strategies aimed at increasing the rate of return to work warrant attention.

### Quality of Life

The multiple linear regression model for health-related QoL (EQ-5D index) revealed that a higher number of unmet needs was significantly associated with a lower QoL, in addition to age, mRS score, and previous stroke. These results are consistent with the findings of previous studies. In a study by Kim et al., an increased number of unmet needs, older age, and mRS scores were significantly associated with a lower health-related QoL as measured by the EQ-5D index (8). In a study by Lehnerer et al., stroke survivors with unmet social needs had significantly lower EQ-5D index values than those with met needs (20). A higher number of unmet needs can lead to lower QoL in stroke survivors. Therefore, LTC systems that monitor and address unmet needs are needed to improve the QoL after stroke. In Korea, community-based rehabilitation is provided for LTC of community-dwelling disabled patients (21). Social care systems should systematically evaluate the unmet needs of stroke survivors and take action to enhance their QoL.

### Perceived Unmet Rehabilitation Needs

The observed prevalence of unmet rehabilitation needs was 32.6%, which is consistent with a previously reported range of 15%-33% (7,22–24). In a study by Ullberg et al., female sex, living alone, smoking, atrial fibrillation, diabetes, somnolence, comatose status, prior stroke, and intracranial hemorrhage or unspecified stroke were all associated with an increased OR for unmet rehabilitation needs (7). Tistad et al. reported that strength at 3 months post-stroke, hand function, and poor self-rated recovery at 12 months were significantly associated with unfulfilled rehabilitation needs at 12 months (23). Similarly, the present study showed that perceived unmet rehabilitation needs were significantly higher in stroke survivors with lower functional status and problems in domains related to QoL. Older stroke patients with lower educational levels also reported more unmet rehabilitation needs. This association of older age and lower educational levels with increased perceived unmet rehabilitation needs may be linked to the generally poorer health outcomes in these stroke survivors (25). Furthermore, older stroke survivors with lower educational and functional statuses may have more difficulties overcoming barriers to community-based rehabilitation, including those related to costs, accessibility, and knowledge (26). These perceived unmet rehabilitation needs in stroke patients with low functional status and multiple comorbidities can be addressed by more individualized rehabilitation plans with realistic goal-setting along with efforts to provide adequate rehabilitation for patient groups facing challenges in accessing medical rehabilitation services after stroke.

### Study Limitations

This study had some limitations. First, of the 9,204 eligible patients, 1,002 were selected for the study, leading to a risk of non-response bias. Comparative analysis of survey responders with non-responders and the whole group revealed that responders were significantly younger, had a shorter time since stroke onset, lower stroke severity, lower mRS score at discharge, a higher proportion of home discharge, and fewer comorbidities (Supplementary table 4). These results may differ for survivors with more severe stroke. However, our findings indicated that even mild stroke survivors have significant unmet needs. Moreover, severe stroke is associated with a higher incidence of unmet needs. Thus, unmet needs may have been underestimated in this study. Second, our data were obtained from four centers and may not be representative of the nationwide population of stroke survivors. Nonetheless, these centers are RCCVCs that play a significant role in the provision of stroke care. Thus, they may be considered representative institutions in their respective areas. Finally, the assessment of unmet needs was based on self-reports, and factors other than stroke may have influenced the reports on unmet needs.

### Conclusions

Long-term unmet needs are prevalent in stroke survivors. Higher unmet needs were associated with a lower QoL and increased odds of reporting unmet rehabilitation needs. The likelihood of perceived unmet rehabilitation needs increased with higher stroke severity, lower functional status, hyperlipidemia, and a higher number of unmet needs. Further research is needed to investigate strategies for addressing these subjective unmet needs with the aim of improving the long-term QoL of stroke survivors.

## Data Availability

The pseudonymized data that support the findings of this study are available from the corresponding author, Pf. Nam-Jong Paik, or the IRB of Seoul National University Bundang Hospital, upon reasonable request, subsequent approval from the local IRB, and completion of a legal data sharing agreement.

## Abbreviations

CI: Confidence interval
EQ-5D-3L: EuroQoL 5-dimension, 3-level
LTC: Long-term care
LUNS: Longer-term Unmet Needs after Stroke
mRS: modified Rankin Scale
NDRS: National Disability Registration System
NIHSS: National Institutes of Health Stroke Scale
OR: Odds ratio
QoL: Quality of life
RCCVC: Regional Cardiocerebrovascular Disease Center
SD: Standard deviation
TIA: Transient ischemic attack

